# Early Side Effects after Administration of the 1st Dose of Oxford-AstraZeneca Vaccine

**DOI:** 10.1101/2022.08.04.22278415

**Authors:** Pramod Singh, Abdul Rafae Faisal, M. Hassaan Shah, Ahmad Saeed, Hadia Younas, Usamah Saeed Butt, Sudip Pudasaini, Abdul Rafay Pasha, Umair Rehman

## Abstract

Vaccines have played a central role in minimizing new infections, the rate of hospitalizations, and the overall burden on the health sector. Fear of side-effects is the biggest and commonest reason for avoiding getting vaccinated. It is, therefore, essential to maintain the clarity and consistency of message, to support and encourage people to get vaccinated. This study aims to contribute in that regard, by registering and quantifying the early side-effects of the Oxford-AstraZeneca COVID-19 vaccine in Pakistan. This study employs a non-random cross-sectional design. Data collected from 477 participants using a structured questionnaire was used to investigate the relationship between socio-demographic characteristics and side effect profiles of the participants. Binomial Logistic Regression was used to analyze the data. Odds Ratio (OR) gives the likelihood of having a side effect versus the reference group. Significance level (α) for the probability value (p-value) is set at 0.05. Fever (30.19%) was the most commonly experienced side effect, followed closely by fatigue (22.01%). 71.11% of those with fever experienced low grade fever (99-100F) while 62.69% of body aches experienced were moderate in intensity (Grades 4-6). In general, younger people are significantly more likely (p=0.023) to experience side effects (OR^-1^ = 1.023: interpreted as 1.023 times increase per unit decrease in age). Similarly, they are more likely (p= 0.029) to have a headache (OR^-1^ =1.039). Also, they are more likely (p= 0.007) to have a body ache (OR^-1^ =1.038). The Oxford-AstraZeneca COVID-19 vaccine side-effects seem to be more prevalent among younger age groups, which points to increased vaccine safety among older individuals that are usually more susceptible to severe COVID-19 infection. In addition, we found a substantially reduced number of side-effects, as compared to the clinical trials, which is an encouraging indicator for vaccine safety.

## Introduction

Since the COVID-19 pandemic started in early 2020, the world has undergone drastic and disruptive changes. The wave upon wave of new infections, constantly having to follow restrictions and fears of the deadly disease has left enduring scars on the public psyche [1]. Not only this, but the pandemic has led to trillions of dollars of loss to the world economy [2].

For this pandemic to end, vaccines have played a central role in minimizing new infections, the rate of hospitalizations and the overall burden on the health sector. A large percentage of the world’s population has to be vaccinated in a short period of time to achieve the herd immunity required to stop the propagation of the Coronavirus [3].

These vaccines have varying mechanisms of actions. Some are mRNA vaccines, like Pfizer-BioNtech and Moderna COVID-19 vaccines, while others, like the Janssen shot, are non-replicating viral vector vaccines. Moreover, the Oxford-AstraZeneca vaccine is an Adenovirus vaccine and Sputnik V is a recombinant Adenovirus vaccine. Many others are inactivated vaccines like Sinopharm and Sinovac vaccines [4, 5].

Side effects have been the cause of concern for many and have led to delay in getting vaccinated. This delay in vaccine acceptance, vaccine hesitancy, is turning out to be an emerging public health problem. [6, 7] According to multiple studies, fear of side-effects is the biggest and commonest reason for avoiding getting vaccinated [8, 9]. It is, therefore, essential to maintain the clarity and consistency of message, to support and encourage people to get vaccinated.

Pakistan started its vaccination campaign with Sinopharm, Sinovac, CanSino and Sputnik V [10]. This effort received public backlash from some quarters. Keeping in mind the dire need of the rapid vaccination of the masses, it is important to maintain the public’s trust, which can only be based on adequate statistical data regarding the safety of the vaccine. This study aims to contribute in that regard, by registering and quantifying the early side-effects of the Oxford-AstraZeneca COVID-19 vaccine in Pakistan.

## Materials and Methods

The study was conducted in July 2021. All individuals presenting at the vaccination site that were within government-approved age groups (at the time of data collection, 40 years and above) were considered for the study.

Individuals with acute febrile illness (last 72 hours), history of COVID-19 infection (positive PCR), prior COVID-19 vaccination., history of foreign travel during the past 3 months, receiving any investigational drug for COVID-19 (including for prophylaxis) during the last 30 days, receiving systemic immunoglobulins, monoclonal antibodies and/or convalescent serum during the past 4 months, significant or severe co-morbidities [Diabetes mellitus, hypertension, chronic pulmonary disease, asthma, active smoker or vapers (during the past year), BMI> 30 Kg/m2, history of anaphylaxis or urticaria or other adverse effects related to vaccine or its components, severe or uncontrolled heart disease, cancer and any other severe disease, bleeding disorder, known or suspected immunodeficient status [HIV, immunosuppressive drugs (cytotoxins and steroids) and asplenia], blood donation during the last 28 days, known or suspected alcohol or drug dependency, pregnancy or breastfeeding, participating in any research study during the past 28 days were excluded from the study.

This study employs a non-random cross-sectional design. Data collected from 477 participants using a structured questionnaire was used to investigate the relationship between socio-demographic characteristics and side effect profiles of the participants. Only those participants that met the inclusion criteria for this study were selected for follow up. The participants were followed on Day 1, 3, and 7 to gather data regarding side-effects. Data was analyzed descriptively and inferentially by IBM SPSS 24. Binomial Logistic Regression was used to analyze the data. For missing data regarding Educational status, data was imputed using mice package in R. Odds Ratio (OR) gives the likelihood of having a side effect versus the reference group. Significance level (α) for the probability value (p-value) is set at 0.05.

## Results

477 individuals participated in this study. Of them, 51.15% of the respondents belonged to the 40-49 age group, followed by the 50-59 age group which had 31.24% participants. 44.89% had primary education or below. A vast majority, 72.54%, were male. Fever (30.19%) was the most commonly experienced side effect, followed closely by fatigue (22.01%). 71.11% of those with fever experienced low grade fever (99-100F) while 62.69% of body aches experienced were moderate in intensity (Grades 4-6) [Table 1]. In general, younger people are significantly more likely (p=0.023) to experience side effects (OR^-1^ = 1.023: interpreted as 1.023 times increase per unit decrease in age). Similarly, they are more likely (p= 0.029) to have a headache (OR^-1^ =1.039). Also, they are more likely (p= 0.007) to have a body ache (OR^-1^ =1.038). In general, higher educational status reported a higher frequency of side effects (p< 0.05) [Table 2].

**Table 1.** Frequency of side effects.

| Side effects | Frequency | Percentage |
| --- | --- | --- |
| Headache | 21 | 4.4 |
| Flu-like illness | 0 | 0 |
| Pain, swelling, rash, bleeding or redness at the site of injection | 56 | 11.74 |
| Fatigue | 18 | 3.77 |
| Anaphylaxis reaction | 1 | 0.21 |
| Fever | 144 | 30.19 |
| Low Grade [99F -100F] | 64 | 71.11 |
| Moderate [100.1F -103 F] | 21 | 23.33 |
| High grade [103.1 F-106 F] | 5 | 5.56 |
| Undocumented | 54 | - |
| Body ache | 105 | 22.01 |
| Grade (1-3) | 8 | 11.94 |
| Grade (4-6) | 42 | 62.69 |
| Grade (7-9) | 17 | 25.37 |
| Undocumented | 38 | - |

**Table 2.**
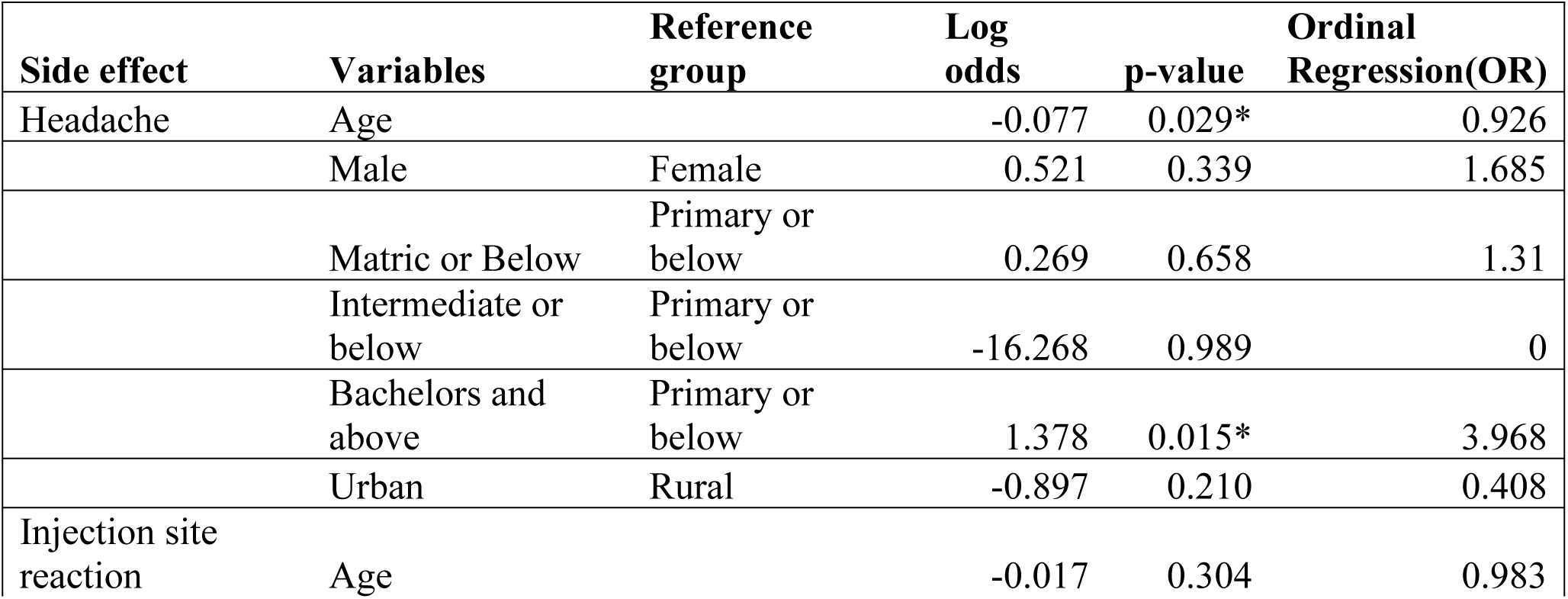
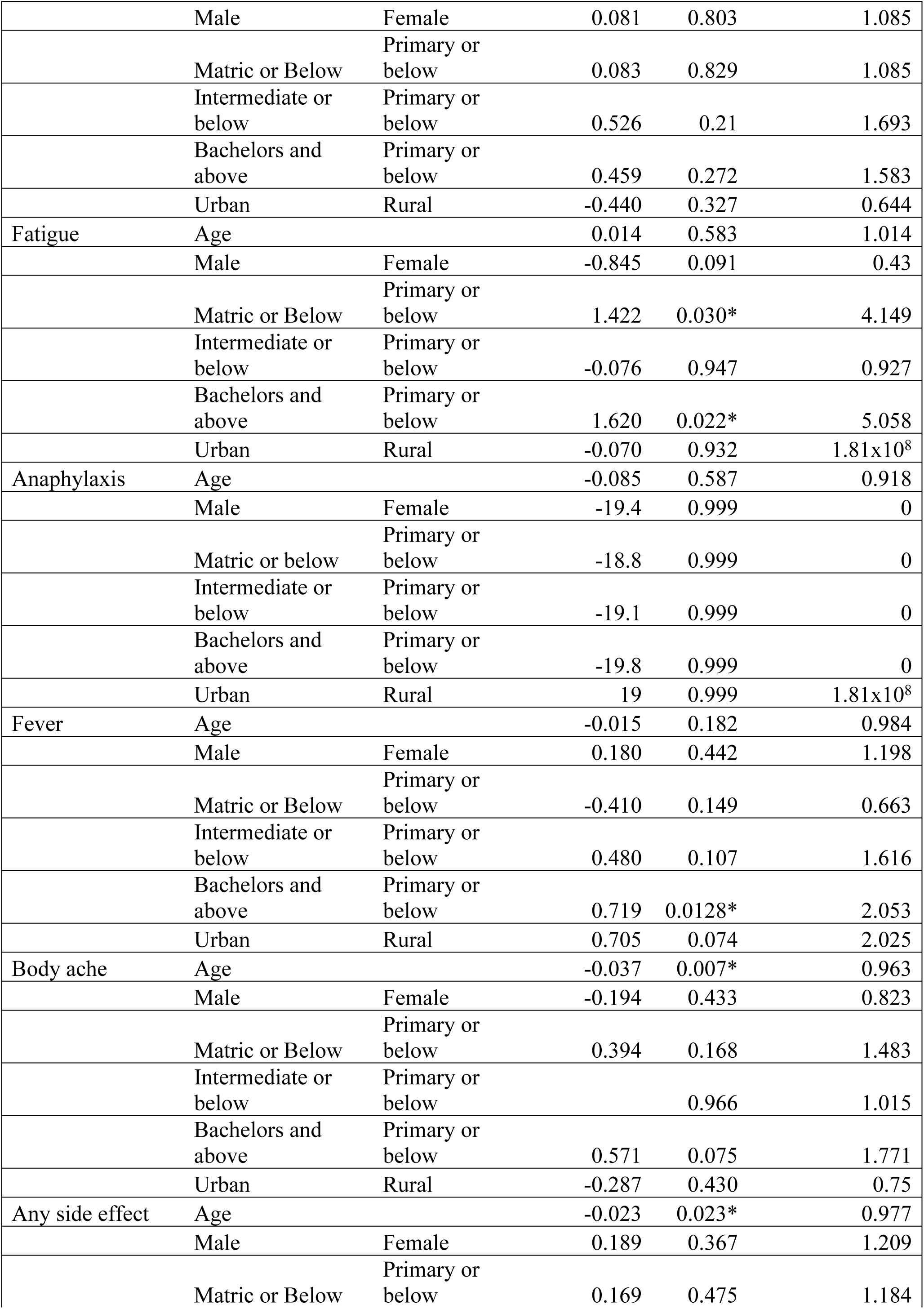
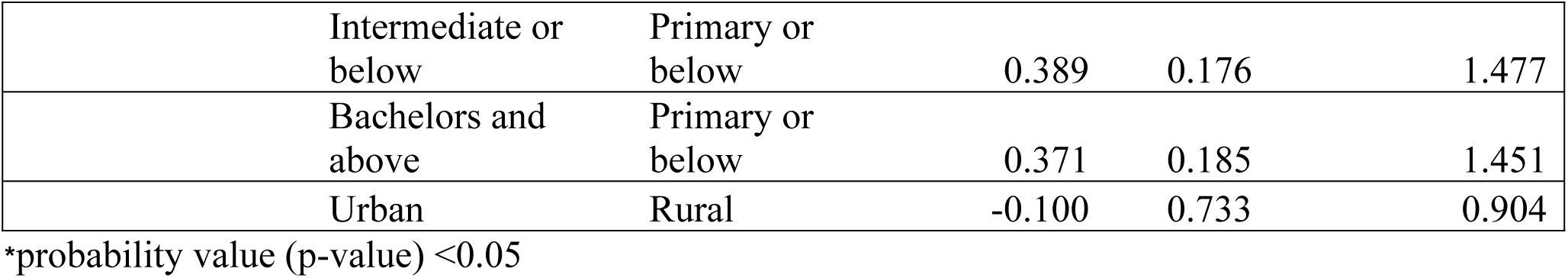
Binary Logistic Regression.

## Discussion

In this study, we attempted to confirm the presence and measure the frequency of occurrence of AstraZeneca vaccine’s early adverse effects in Pakistan. These included headache, body aches, fatigue, fever, injection site reaction, flu-like illness and anaphylaxis. Except for the flu-like illness and anaphylaxis, for all COVID-19 vaccines, these early side effects are, generally speaking, the commonest early COVID-19 vaccine side effects [11]. 49.47% of the respondents in our study had, at least, one of these symptoms.

The side-effects were categorized as local and systemic. Local effects refer to injection site reactions which include pain, swelling, rash, redness or bleeding at the site of vaccination. They were present only in 11.74% of the study population which is in marked contrast to phase 2/3 trials, that showed a considerably higher occurrence across all age groups (88% in the 18–55 years group, 73% in the 56–69 years group, and 61% in 70 years and older group), and phase 1/ 2 trials, conducted in UK, at 67% [12, 13]. It is, however, much closer to a regional study conducted in Bangladesh (35.5%) [14] Most of the local adverse effects occurred on the first and second post-vaccination day, while a few were recorded on the third day. None were reported in the following days. This is in line with the results of the clinical trials [12].

On the other hand, the systemic adverse effects solicited included headache, fever, flu-like illness, body aches, fatigue and anaphylaxis. At least one systemic effect was reported by 42.76% of the study participants, which is considerably lower than the results of the phase 2/ 3 trials (86% in the 18–55 years group, 77% in the 56–69 years group, and 65% in the 70 years and older group) [12].

Fever, at 30.19%, was present in a greater number of individuals than any other side effect. The phase 2/ 3 trials show a similar result with 24% incidence in the 18-55 age and no reports of fever in the 55 or above age group. They are also similar to the results of the Bangladeshi study (20%) mentioned above [12, 14]. Another study conducted on those 50 years old and above reported a lower incidence of 10% [15].

Of those that reported fever, nearly three quarters (71.11%) had only a mild case of it. Another 23.33% reported moderate fever and very few individuals reported incidence of high grade fever. This is in accordance with phase 2/3 trials [12].

Body aches were reported by 22.01% of the participants. This figure is much lower than the phase 1/2 trials (60%) and much closer to a regional study (16.4%) [13, 14]. Body aches were categorized using a numerical scale from 1 to 10. Over half (62.69%) of the people with body aches reported moderate (4 to 6) while a significant number (25.37%) of them also had severe body aches (7 to 10). This agrees with prior reports of mostly moderate systemic side-effects. [16]

Headache was present in a mere 4.40% of the respondents. This is, again, in concurrence with the Bangladeshi study (6.9%) rather than the clinical trial results (68%). Fatigue was reported by 3.77% of the participants, a result similar to the Bangladeshi study, and there was not a single case of flu-like illness [14].

One case of anaphylactic reaction was also registered, which is in accordance with its rare occurrence in similar studies [12, 16]. This hypersensitivity is probably due to polysorbate 80 which is present in adenoviral vector vaccines. These excipients serve to increase absorption, improve solubility, and enhance stability of the whole product [17, 18].

The development and progress of side-effects were assessed through multiple follow-ups during the 7-day time period. All of them appeared on Day 1 and most disappeared by Day 4. A majority of symptoms were present on Day 2 (frequency = 316), followed by Day 1 (219) and Day 3 (122). There was always a dramatic fall off, by Day 4 (13). Rarely did a symptom persist till Day 5 (2).The mean duration of symptoms was (1.9 ± 0.7), which is very similar to reported in a study in Bangladesh (1.9 ± 1.3 day).

The inferential analysis yielded a significant inverse relationship between age and headache and age and body aches. Generally, younger individuals were at a greater risk of exhibiting adverse effects than their older counterparts. This is in accordance with multiple studies [12, 16, 19]. An explanation of this phenomenon lies in the fact that younger individuals form stronger immune responses which naturally lead to a greater number of such side-effects [20]. That may be a reason for the reduced frequencies of side-effects in this study, as the average age was 51.28 years (AstraZeneca vaccine was available only for those who were 40 years old or above, at the time of data collection). Nevertheless, it cannot be the sole cause behind considerable differences in results. This is supported by the fact that some studies have shown that there are far fewer real-world side-effects of these vaccines [14, 19]. Natural variations among different populations also exist and must have a part to play in it.

No significant associations were found for sex or residence. This is in contrast to past studies that found an association between sex and the occurrence of side-effects [17, 19].

Studies on vaccine side-effects cite past COVID-19 infection and co-morbidities as among the risk factors for the emergence of post-vaccination adverse effects (although one study shows an opposite relationship for the latter) [14, 21].

## Conclusion

The Oxford-AstraZeneca COVID-19 vaccine side-effects seem to be more prevalent among younger age groups, which points to increased vaccine safety among older individuals that are usually more susceptible to severe COVID-19 infection. They are not associated with sex and residence. However, education is positively associated with them, which might indicate a nocebo effect among the educated resulting from higher awareness and knowledge about side-effects. Finally, we found a substantially reduced number of side-effects, as compared to the clinical trials, which is an encouraging indicator for vaccine safety.

We enacted stringent and expansive exclusion criteria to eliminate nearly all chronic illnesses and co-morbidities that may influence the results. Similarly, we did not include individuals with past COVID-19 infection (positive PCR test in the past). This was done in the interest of reducing all possible risk factors, which may inflate the figures, to a minimum and to increase the confidence in the results generated. While it is a fact that increasing age is inversely correlated with the development of said side-effects, as mentioned above, the responsibility for the considerably lower frequency of their occurrence cannot be laid solely at the feet of higher median age, in this study. Further independent, prospective studies are required to detect all the factors at play.

## Data Availability

URL: https://osf.io/ndf5r/

## Acknowledgements

None.

## Limitations

As the follow-ups were done via phone calls rather than in-person (owing to pandemic-induced restrictions), the data is subject to reporting and recall bias. At any rate, 3 equally spaced follow-ups, which allowed for cross and secondary questioning, kept such issues to a minimum. No serological testing was included, which may give an idea about the unknown risk factors.

